# Health systems performance for hypertension control using a cascade of care approach in South Africa, 2008-2017

**DOI:** 10.1101/2021.09.13.21251870

**Authors:** Mariet Benade, Sridevi K. Prasad, Zandile Mchiza, Lily D. Yan, Alana T. Brennan, Justine Davies, Nikkil Sudharsanan, Jennifer Manne-Goehler, Matthew P. Fox, Jacob Bor, Sydney B. Rosen, Andrew C. Stokes

**Affiliations:** Department of Global Health, Boston University School of Public Health, Boston, Massachusetts, USA; School of Public Health, University of the Western Cape, Bellville, South Africa; Department of Medicine, Weill Cornell Medicine, New York, New York, USA; Department of Epidemiology, Boston University School of Public Health. Department of Global Health, Boston University School of Public Health, Boston, Massachusetts, USA; Health Economics and Epidemiology Research Office, Department of Internal Medicine, School of Clinical Medicine, Faculty of Health Sciences, University of the Witwatersrand, Johannesburg, South Africa; Department of Global Health, University of Birmingham, Birmingham, United Kingdom; Heidelberg Institute for Global Health, Heidelberg University, Heidelberg, Germany; Division of Infectious Diseases, Massachusetts General Hospital, Boston, MA; Department of Global Health, School of Public Health, Boston University, Boston, Massachusetts, USA

## Abstract

**Background:** Hypertension is a major contributor to global morbidity and mortality. In South Africa, the government has employed a whole systems approach to address the growing burden of non-communicable diseases. We used a novel incident care cascade approach to measure changes in the South African health system’s ability to manage hypertension between 2008 and 2017.

**Methods:** We used data from Waves 1-5 of the National Income Dynamics Study (NIDS) to estimate trends in the hypertension care cascade and unmet treatment need across four successive cohorts with incident hypertension. We used a negative binomial regression to identify factors that may predict higher rates of hypertension control, controlling for socio-demographic and healthcare factors. The largest cascade attrition occurred prior to diagnosis.

**Results:** In 2011, 19·6% (95%CI 14·2, 26·2) of individuals with incident hypertension were diagnosed, 15·4% (95%CI 10·8, 21·4) were on treatment and 7.1% had controlled blood pressure. By 2017, the proportion of individuals with diagnosed incident hypertension had increased to 24·4% (95%CI 15·9, 35·4) with increases in treatment (23·3%, 95%CI 15·0, 34·3) and control (22·1%, 95%CI 14·1, 33·.0) were also observed, translating to a decrease in unmet need from 92·9% in 2011 to 77·9% in 2017. Multivariable regression showed that participants with incident hypertension in 2017 were 3·01 (95%CI 1·77, 5·13) times more likely to have a controlled blood pressure compared to those in 2011.

**Conclusions:** The proportion of people with incident hypertension who successfully progressed to controlled blood pressure tripled between 2011 and 2017 in South Africa. Despite these improvements, a low absolute proportion of the population were able to control their blood pressure and a high burden of unmet need remains.

**Summary Box:** *What is already known:*

- Prevalent cascades provide insight to where losses in care cascades occur.
- While mostly used in the management of HIV, recently they have also been adopted in studying the management of non-communicable diseases on a population level.
- Prevalent hypertension cascades in South Africa showed a high burden of unmet need, with the biggest losses where lost between disease development and diagnosis.

*What are the new findings:*

- Incident hypertension cascades improved from 2008 to 2017 in South Africa.

*What do the new findings imply:*

- Incident cascades provide an improved means to measure changes in management cascades as this allows us to distinguish between historical and current health system performance.
- Our data show that while substantial improvements in the care cascade occurred between 2008 and 2017, a large burden of unmet need remains.

## Introduction

Hypertension is a major contributor to global morbidity and mortality, resulting in an estimated 10·4 million deaths in 2018 and 218 million disability adjusted life years (DALYs) in 2017 through its effects on cardiovascular disease and stroke.[1] Despite the availability of numerous inexpensive evidence-based interventions such as lifestyle modification and pharmacotherapy, prevalence and complication rates have continued to rise globally.[2–4] This trend affects low- and middle-income countries (LMIC’s) disproportionately. More than a fifth (21%) of all deaths in LMIC’s are attributed to hypertension and prevalence there has increased by 7·7% between 2000 and 2010, while it has decreased by 2.6% in high-income countries.[5,6]

In South Africa, the prevalence of hypertension was 46% in women and 44% in men in 2016.[7] Among people living with hypertension, the proportion with controlled hypertension was found to be only 20% in women and 13% in men that year, suggesting that substantial gaps in coverage and quality of hypertension care remain.[7] Previous work has found that low rates of diagnosis and treatment among those with hypertension create a high burden of unmet need for hypertension management.[7,8] A cost-effectiveness analysis in 2019 concluded that increasing the diagnosis and timeliness of treatment in South Africa would substantially reduce DALYs attributed to hypertension while also cutting health care costs related to complications of hypertension, potentially saving the South African health care system $24, 902 per DALY averted.[9]

Although the South African government has undertaken numerous health systems reforms in recent years with the aim of strengthening management of chronic non-communicable diseases, including improved guidelines and the *Strategic Plan for the Prevention and Control of Non-Communicable Diseases 2013-2017*, the extent to which new policies have translated to population-wide improvements in hypertension control remains unclear.[10–12]

One approach to tracking the appropriateness of health systems interventions are care cascades. Care cascades can identify the specific stages of care at which interventions should be focused to improve chronic disease control, and monitor progress over time, as suggested by the 2018 Lancet Global Health Systems Strengthening Commission.[13] By measuring how individuals move through a health care system screening and diagnosis to treatment and, ultimately, control, care cascades reflect patient management success in health systems and capture which components of health service delivery require the most improvement.[14,15]

Despite their potential value, current care cascade analyses for hypertension are generally based on prevalent samples assessed at a single point in time, and thus cannot capture time trends or distinguish between historical and current health system weaknesses.[16] More nuanced health systems planning requires repeated assessment of cohorts with new onset of disease to allow inferences of trends in care cascades to attributes of the contemporary health care system, as has recently been advocated for in HIV cascades.[17]

In the present study, we evaluate changes in the cascade of care for hypertension in South Africa over the period 2008-2017 using data from the National Income Dynamics Study (NIDS). We develop a novel longitudinal approach to analyzing hypertension care cascades in which we examine successive cohorts of people who newly develop hypertension over the defined period. Incident cascades offer a clearer depiction of health systems performance at specific points in time, as it allows for a consideration of movement of new cases through the cascade in a specified time period. Using data from all five waves of NIDS, we use incident cascades to assess current health systems performance and the potential effects of policies introduced over the study period.[18–22]

## Methods

### Design and setting

This study uses data from the first through the fifth waves of the National Income Dynamics Study (NIDS).[18–22] NIDS is a nationally representative panel survey that was implemented by the South African Labour and Development Research Unit at the University of Cape Town’s School of Economics to track changes in the structure and living conditions of South African households and the living conditions and the well-being of individual household members over time.

The first wave of data was collected in 2008, with subsequent waves collecting data from the same households and their individual members in 2011, 2013, 2015 and 2017. The initial sample population consisted of 28,000 individuals from 7,300 households across South Africa. Using a stratified, two-stage cluster sample design, households were first stratified using 53 district councils with 400 primary sampling units, then randomly selected within each stratum.[23] Four subsequent waves collected data from the same households and their individual members in 2011, 2013, 2015 and 2017.

Data were collected by trained interviewers during face-to-face interviews conducted in the participant’s preferred language. Similar questions were asked of participants at each wave of the survey. Questions relevant to this study included age and known illnesses (asked in free form) and education level, and ethnicity (asked with a list of options). Blood pressure measurements were conducted at the end of each interview, with two measurements taken at five-minute intervals.

### Participants

To create our sample population, we first identified those who were sampled in NIDS wave one. We then excluded all participants who were lost to follow up and thus not successfully interviewed in all four subsequent waves (N=10,013). Following that, we restricted the sample to participants that were aged 25-79 years at wave one (N=4,884) and then excluded all those with missing hypertension, diagnosis, treatment or control data (N=1992). We were left with a sample of 2892, from which we identified those who met the criteria for incident hypertension described below. Our final sample comprised 747 incident hypertension cases that occurred any time after wave one (i.e. discerned in waves two through five) (Supplementary Figure 1). Attrition rates were highest among White (50·07%-62·69%) and Indian/Asian (36·44%-44·82%) participants and participants in high income categories.[24] Average attrition rates between waves ranged from 14·01% to 21·93%.

### Incident Cohort Design

To create our incident cohorts, we identified people who met our incident hypertension case definition at each wave of the NIDS study. We defined an individual as newly hypertensive if they had high blood pressure (defined below) in wave two, three, four or five, and had never reported being hypertensive in any previous waves. Using the newly hypertensive cases in each wave, we created incident cohorts of new cases of hypertension across all 4 waves (Supplementary Figure 2).

### Care Cascade

Care cascades using incident cohorts from all waves were created to measure the percentage of the incident population that successfully progressed to each phase of the cascade, which enabled us to calculate losses at each phase and overall burden of unmet need. We defined a care cascade for hypertension with four stages: (1) hypertensive, (2) diagnosed, (3) treated and (4) controlled. Individual participants’ entry into each subsequent stage of the cascade was contingent on them having reached the previous stage (Figure 1).

**Figure 1:**
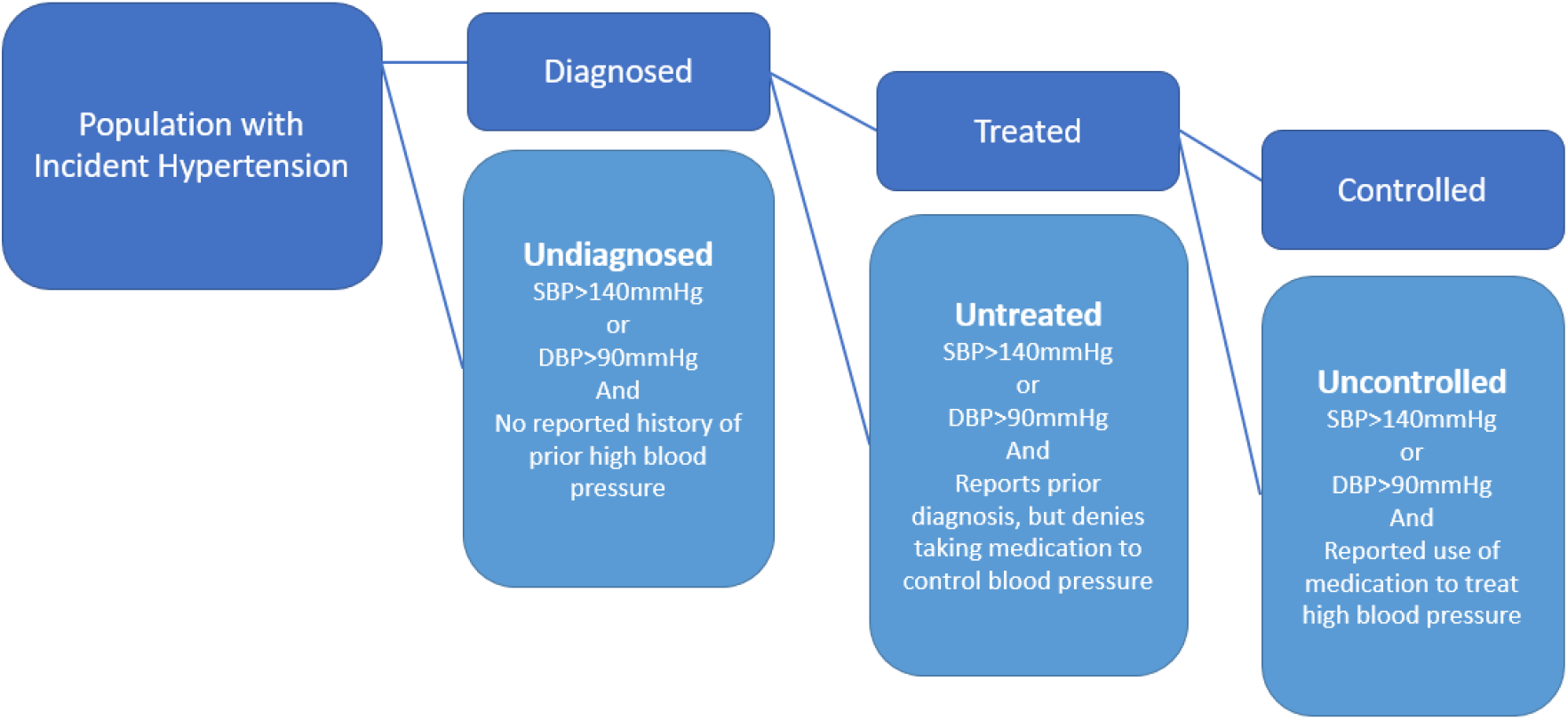
Conceptual Design Figure SBP, Systolic Blood Pressure DBP, Diastolic Blood Pressure

Using the current South African guidelines, we defined an individual as hypertensive (stage 1) if their average systolic blood pressure was equal or higher than 140 mmHg, their average diastolic blood pressure equal or higher than 90mmHg, or they self-reported taking anti-hypertensive treatment. We defined participants as diagnosed (stage 2) if they answered “yes” to the question “Have you ever been told by a doctor, nurse or health care professional that you have high blood pressure?”.

Participants were determined to be “treated” (stage 3) if they reported being diagnosed and that they were taking medication for high blood pressure and “untreated” if they were not. Finally, participants were determined to be “controlled” (stage 4) if they had been diagnosed, were on treatment and had a blood pressure less than 140 mmHg systolic and 90 mmHg diastolic. Participants who answered “yes” to the treatment question but had a blood pressure greater than 140 mmHg systolic or 90 mmHg diastolic were determined to be “uncontrolled”.

We defined “unmet need” as the sum of the proportion of the sample who remain undiagnosed, untreated or uncontrolled.

### Covariates

We included variables on sex, age group, race, education, province, urban or rural residence, medical aid (a form of private health insurance), year of survey, income, known comorbidities, and whether a participant had seen a health care worker in the past year. Age groups were 35-44 years, 45-59 years, and 60-79 years. Race was classified according to the official groupings used by the South Africa Bureau of Statistics. For education we created a dichotomous variable that distinguished between those who completed schooling up to grade nine, which is compulsory in South Africa, and those who completed schooling beyond the ninth grade. Year of survey was derived from the survey wave in which the participant met the criteria of an incident hypertension case. We created a new ordinal variable that assigned participants to an income quartile based on their total household income. We considered patients to have a known comorbidity if they responded “yes” to if they had ever been told that they had diabetes, stroke, heart disease or cancer.

### Statistical Analyses

Hypertension estimates were age-standardized to the population of South Africa using the 2017 census report.[25] We calculated incidence rate ratios using a multivariable negative binomial regression across all waves on the outcomes of being diagnosed, treated or controlled. The main variable of interest was survey wave, as this allowed us to assess changes over time. The model was adjusted for the covariates described above. We generated predicted probability using marginal standardization while controlling for the same covariates. To account for the survey design, each incident cohort was weighted by that wave’s sample weights.[23] All analyses were performed in Stata 16 (StataCorp, College Station, TX).

## Results

Our final sample size for each incident cohort was 335 for wave two, 189 for wave three, 120 for wave four, and 103 for wave five. The proportion of women in each incident cohort decreased over time, from 63·6% in wave two to 46·0% in wave five. There were minor changes in racial composition across the waves, with an overall increase in the proportion of participants of African race from 79·3% in wave two to 85·0% in wave five. Participants identifying as coloured decreased from 11·1% to 7·2% and those of Asian/Indian descent remained stable (7·2% in wave two and 7·5% in wave five) (Table 1).

**Table 1.**
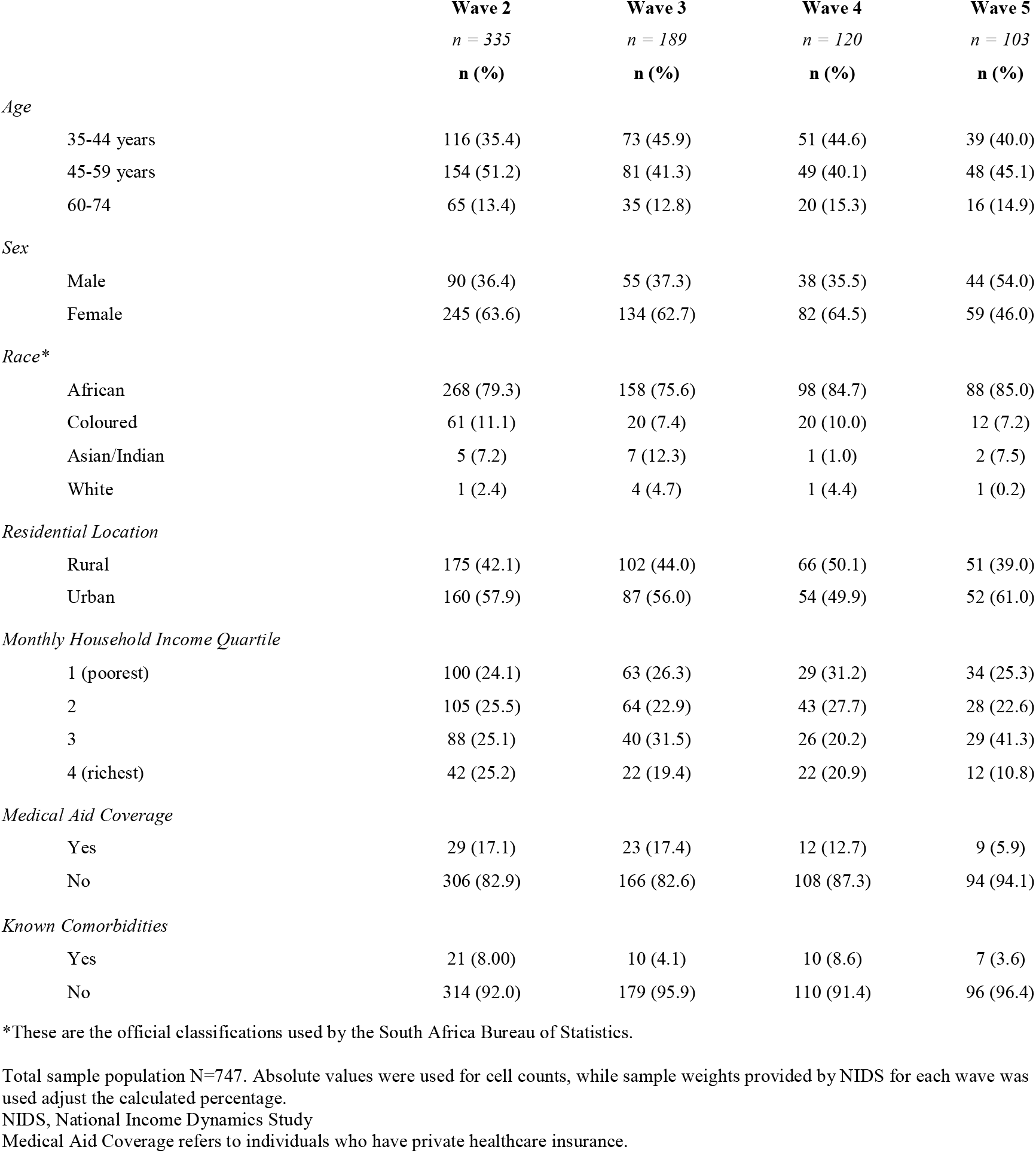
Characteristics of NIDS Incident Hypertension Cohorts, 2008 – 2017, N=747

### Trends in the incident hypertension cascade

Figure 2 shows the hypertension care cascades for each of the four incident cohorts. The proportion of those who were hypertensive who were found to be diagnosed increased from 19·6% (95% CI: 14·2-26·2%) in wave two, to 24·4% (95% CI: 15·9-35·5%) in wave five, after decreasing to 17·5% (95% CI: 10·1-28·4%) in wave three and improving again in wave four to 18·8% (95% CI: 11·5-29·2%). The proportion of those who were hypertensive that were diagnosed and treated also increased from wave two to five (W2: 15·4%, W3: 17·4%, W4: 13·9%, W5: 23·3%), though there were minor reductions in the proportion of those who were on treatment in wave three and four. The proportion of the incident hypertensive population that successfully moved through the cascade from diagnosis to hypertension control increased steadily over time, from 7·1% (95% CI: 4·1-12·0%) in wave two, 11·4% (95% CI: 5·5-22·1%) in wave three, 11·8% (95% CI: 6·1-21·6%) in wave four to 22.1% (95% CI: 14·1-33·0%) in wave five.

**Figure 2.**
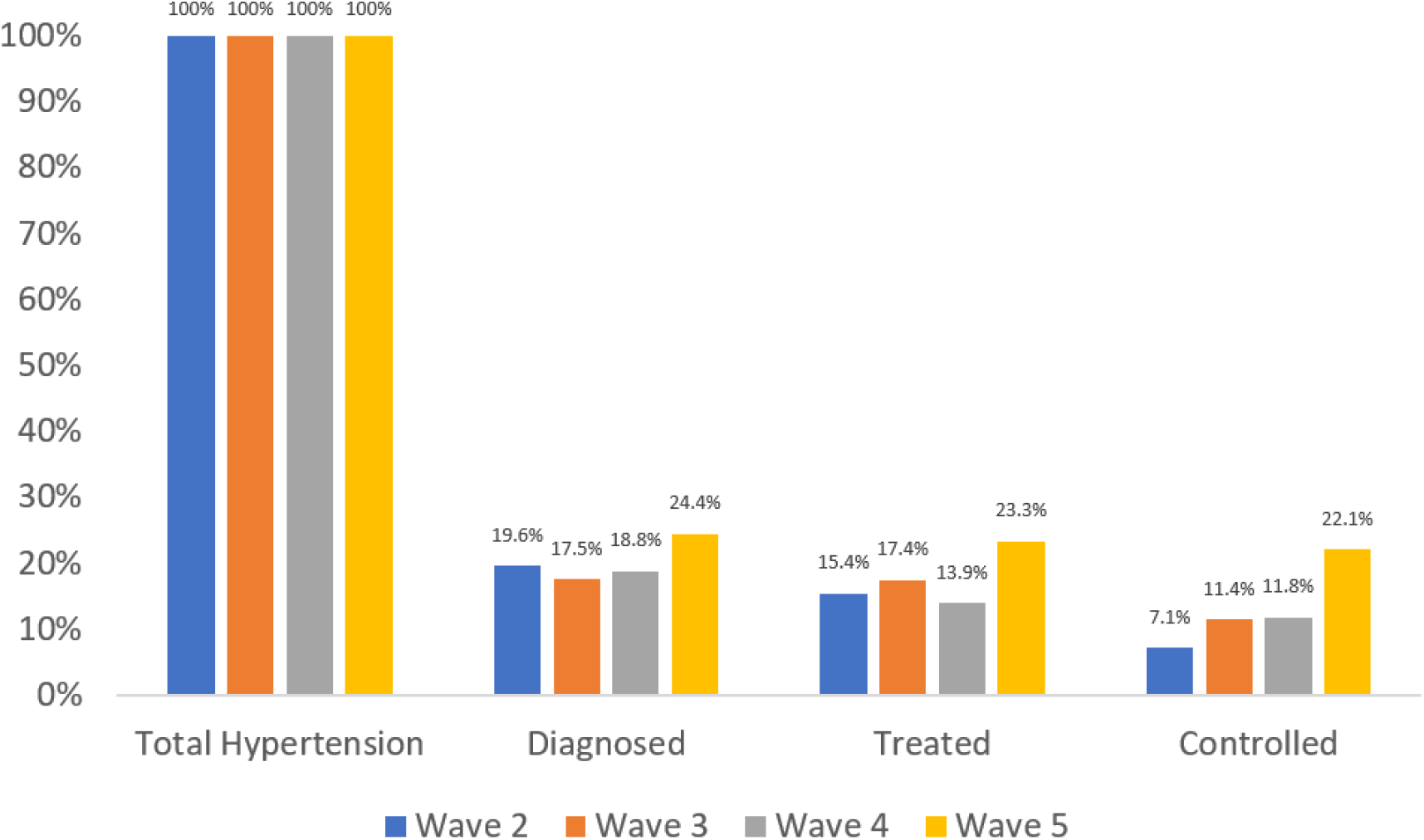
Incident Hypertension Care Cascades, South Africa 2008-2017

The burden of unmet need for hypertension care was estimated to be 92·9% for the wave two incident cohort. In the wave two cohort, 19·6% of new cases of hypertension reported being diagnosed prior to the study interview. Of those who were diagnosed, 78·6% reported taking anti-hypertensive medication. Of those on treatment, 46·1% had a normal blood pressure. This translates to 7·1% of all new hypertension cases in wave two being controlled, leading to a burden of unmet need of 92·9%. Because of the improvements in each stage over time noted above, this burden of unmet need fell substantially to 77·9% in wave 5.

Among newly hypertensive patients across all five waves, 80·59% were undiagnosed (Table 2). An estimated 2·07% were diagnosed, 6·25 were treated and 11·07% were controlled. Overall, the incidence of hypertension decrease with each wave, while the percentage of participants who progressed to being controlled increased. Despite lower incidence (7·04% vs 8·47%), women (15·71%,)were more likely to have their blood controlled than men (4·62%). When considering income quartiles, those in the highest quartile had both the lowest incidence (5·80%) and the highest proportion of those with hypertension be controlled (21·13%).

**Table 2.**
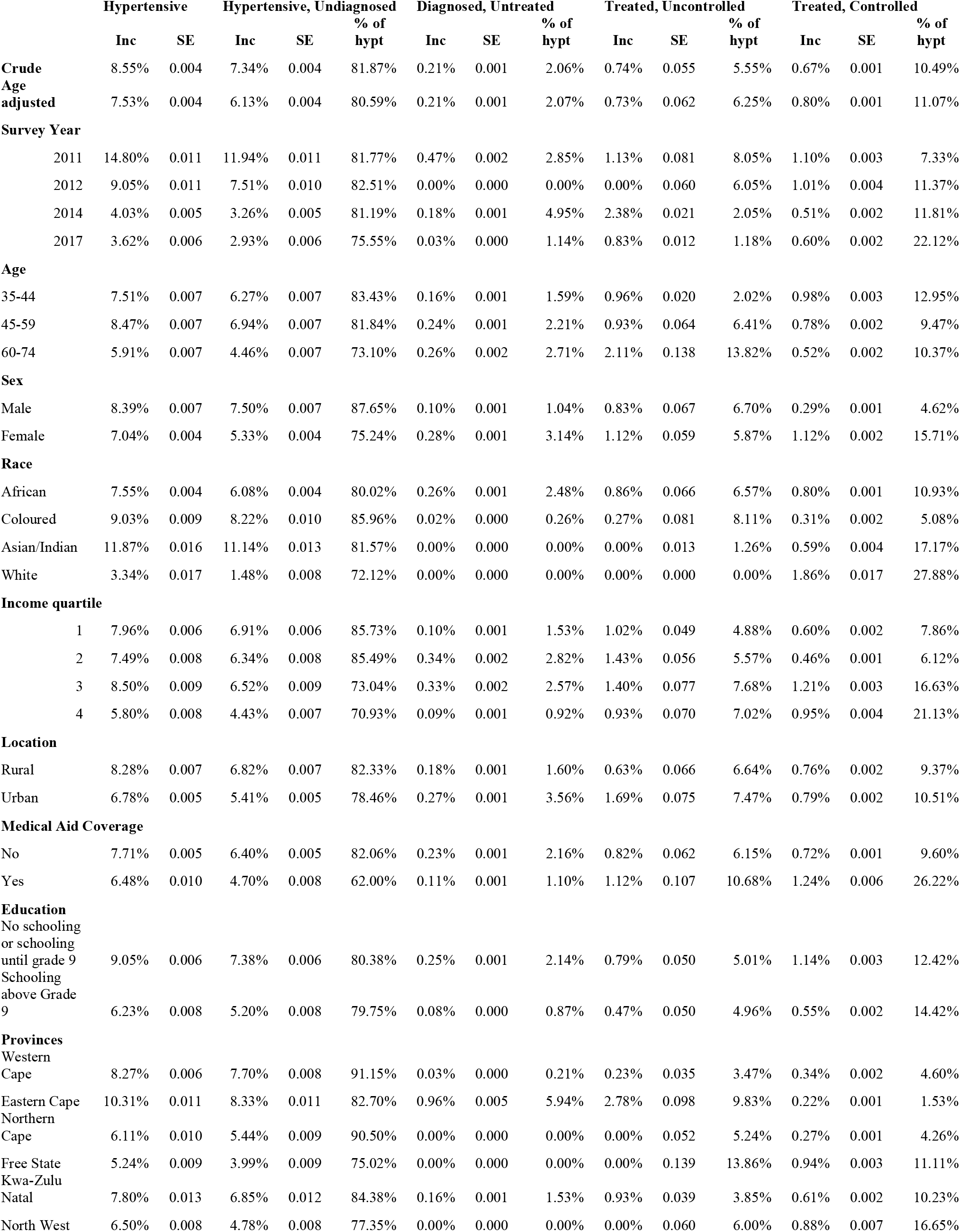
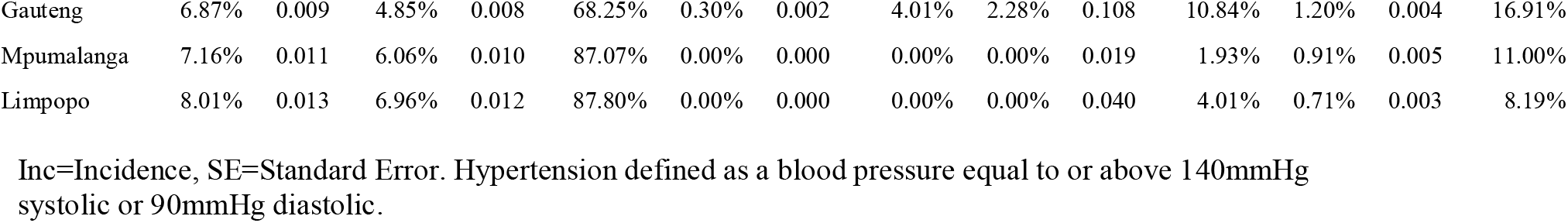
Hypertension incidence, diagnosis, treatment and control, South African Adults aged 35-74, 2010-2017

|  | Hypertensive |  | Hypertensive, Undiagnosed<br>% of |  |  | Diagnosed, Untreated<br>% of |  |  | Treated, Uncontrolled<br>% of |  |  | Treated, Controlled<br>% of |  |  |
| --- | --- | --- | --- | --- | --- | --- | --- | --- | --- | --- | --- | --- | --- | --- |
|  | Inc | SE | Inc | SE | hypt | Inc | SE | hypt | Inc | SE | hypt | Inc | SE | hypt |
| **Crude** | 8.55% | 0.004 | 7.34% | 0.004 | 81.87% | 0.21% | 0.001 | 2.06% | 0.74% | 0.055 | 5.55% | 0.67% | 0.001 | 10.49% |
| **Age adjusted** | 7.53% | 0.004 | 6.13% | 0.004 | 80.59% | 0.21% | 0.001 | 2.07% | 0.73% | 0.062 | 6.25% | 0.80% | 0.001 | 11.07% |
| **Survey Year** |  |  |  |  |  |  |  |  |  |  |  |  |  |  |
| 2011 | 14.80% | 0.011 | 11.94% | 0.011 | 81.77% | 0.47% | 0.002 | 2.85% | 1.13% | 0.081 | 8.05% | 1.10% | 0.003 | 7.33% |
| 2012 | 9.05% | 0.011 | 7.51% | 0.010 | 82.51% | 0.00% | 0.000 | 0.00% | 0.00% | 0.060 | 6.05% | 1.01% | 0.004 | 11.37% |
| 2014 | 4.03% | 0.005 | 3.26% | 0.005 | 81.19% | 0.18% | 0.001 | 4.95% | 2.38% | 0.021 | 2.05% | 0.51% | 0.002 | 11.81% |
| 2017 | 3.62% | 0.006 | 2.93% | 0.006 | 75.55% | 0.03% | 0.000 | 1.14% | 0.83% | 0.012 | 1.18% | 0.60% | 0.002 | 22.12% |
| **Age** |  |  |  |  |  |  |  |  |  |  |  |  |  |  |
| 35-44 | 7.51% | 0.007 | 6.27% | 0.007 | 83.43% | 0.16% | 0.001 | 1.59% | 0.96% | 0.020 | 2.02% | 0.98% | 0.003 | 12.95% |
| 45-59 | 8.47% | 0.007 | 6.94% | 0.007 | 81.84% | 0.24% | 0.001 | 2.21% | 0.93% | 0.064 | 6.41% | 0.78% | 0.002 | 9.47% |
| 60-74 | 5.91% | 0.007 | 4.46% | 0.007 | 73.10% | 0.26% | 0.002 | 2.71% | 2.11% | 0.138 | 13.82% | 0.52% | 0.002 | 10.37% |
| **Sex** |  |  |  |  |  |  |  |  |  |  |  |  |  |  |
| Male | 8.39% | 0.007 | 7.50% | 0.007 | 87.65% | 0.10% | 0.001 | 1.04% | 0.83% | 0.067 | 6.70% | 0.29% | 0.001 | 4.62% |
| Female | 7.04% | 0.004 | 5.33% | 0.004 | 75.24% | 0.28% | 0.001 | 3.14% | 1.12% | 0.059 | 5.87% | 1.12% | 0.002 | 15.71% |
| **Race** |  |  |  |  |  |  |  |  |  |  |  |  |  |  |
| African | 7.55% | 0.004 | 6.08% | 0.004 | 80.02% | 0.26% | 0.001 | 2.48% | 0.86% | 0.066 | 6.57% | 0.80% | 0.001 | 10.93% |
| Coloured | 9.03% | 0.009 | 8.22% | 0.010 | 85.96% | 0.02% | 0.000 | 0.26% | 0.27% | 0.081 | 8.11% | 0.31% | 0.002 | 5.08% |
| Asian/Indian | 11.87% | 0.016 | 11.14% | 0.013 | 81.57% | 0.00% | 0.000 | 0.00% | 0.00% | 0.013 | 1.26% | 0.59% | 0.004 | 17.17% |
| White | 3.34% | 0.017 | 1.48% | 0.008 | 72.12% | 0.00% | 0.000 | 0.00% | 0.00% | 0.000 | 0.00% | 1.86% | 0.017 | 27.88% |
| **Income quartile** |  |  |  |  |  |  |  |  |  |  |  |  |  |  |
| 1 | 7.96% | 0.006 | 6.91% | 0.006 | 85.73% | 0.10% | 0.001 | 1.53% | 1.02% | 0.049 | 4.88% | 0.60% | 0.002 | 7.86% |
| 2 | 7.49% | 0.008 | 6.34% | 0.008 | 85.49% | 0.34% | 0.002 | 2.82% | 1.43% | 0.056 | 5.57% | 0.46% | 0.001 | 6.12% |
| 3 | 8.50% | 0.009 | 6.52% | 0.009 | 73.04% | 0.33% | 0.002 | 2.57% | 1.40% | 0.077 | 7.68% | 1.21% | 0.003 | 16.63% |
| 4 | 5.80% | 0.008 | 4.43% | 0.007 | 70.93% | 0.09% | 0.001 | 0.92% | 0.93% | 0.070 | 7.02% | 0.95% | 0.004 | 21.13% |
| **Location** |  |  |  |  |  |  |  |  |  |  |  |  |  |  |
| Rural | 8.28% | 0.007 | 6.82% | 0.007 | 82.33% | 0.18% | 0.001 | 1.60% | 0.63% | 0.066 | 6.64% | 0.76% | 0.002 | 9.37% |
| Urban | 6.78% | 0.005 | 5.41% | 0.005 | 78.46% | 0.27% | 0.001 | 3.56% | 1.69% | 0.075 | 7.47% | 0.79% | 0.002 | 10.51% |
| **Medical Aid Coverage** |  |  |  |  |  |  |  |  |  |  |  |  |  |  |
| No | 7.71% | 0.005 | 6.40% | 0.005 | 82.06% | 0.23% | 0.001 | 2.16% | 0.82% | 0.062 | 6.15% | 0.72% | 0.001 | 9.60% |
| Yes | 6.48% | 0.010 | 4.70% | 0.008 | 62.00% | 0.11% | 0.001 | 1.10% | 1.12% | 0.107 | 10.68% | 1.24% | 0.006 | 26.22% |
| **Education** |  |  |  |  |  |  |  |  |  |  |  |  |  |  |
| No schooling or schooling until grade 9 | 9.05% | 0.006 | 7.38% | 0.006 | 80.38% | 0.25% | 0.001 | 2.14% | 0.79% | 0.050 | 5.01% | 1.14% | 0.003 | 12.42% |
| Schooling above Grade 9 | 6.23% | 0.008 | 5.20% | 0.008 | 79.75% | 0.08% | 0.000 | 0.87% | 0.47% | 0.050 | 4.96% | 0.55% | 0.002 | 14.42% |
| **Provinces** |  |  |  |  |  |  |  |  |  |  |  |  |  |  |
| Western Cape | 8.27% | 0.006 | 7.70% | 0.008 | 91.15% | 0.03% | 0.000 | 0.21% | 0.23% | 0.035 | 3.47% | 0.34% | 0.002 | 4.60% |
| Eastern Cape | 10.31% | 0.011 | 8.33% | 0.011 | 82.70% | 0.96% | 0.005 | 5.94% | 2.78% | 0.098 | 9.83% | 0.22% | 0.001 | 1.53% |
| Northern Cape | 6.11% | 0.010 | 5.44% | 0.009 | 90.50% | 0.00% | 0.000 | 0.00% | 0.00% | 0.052 | 5.24% | 0.27% | 0.001 | 4.26% |
| Free State | 5.24% | 0.009 | 3.99% | 0.009 | 75.02% | 0.00% | 0.000 | 0.00% | 0.00% | 0.139 | 13.86% | 0.94% | 0.003 | 11.11% |
| Kwa-Zulu Natal | 7.80% | 0.013 | 6.85% | 0.012 | 84.38% | 0.16% | 0.001 | 1.53% | 0.93% | 0.039 | 3.85% | 0.61% | 0.002 | 10.23% |
| North West | 6.50% | 0.008 | 4.78% | 0.008 | 77.35% | 0.00% | 0.000 | 0.00% | 0.00% | 0.060 | 6.00% | 0.88% | 0.007 | 16.65% |
| Gauteng | 6.87% | 0.009 | 4.85% | 0.008 | 68.25% | 0.30% | 0.002 | 4.01% | 2.28% | 0.108 | 10.84% | 1.20% | 0.004 | 16.91% |
| Mpumalanga | 7.16% | 0.011 | 6.06% | 0.010 | 87.07% | 0.00% | 0.000 | 0.00% | 0.00% | 0.019 | 1.93% | 0.91% | 0.005 | 11.00% |
| Limpopo | 8.01% | 0.013 | 6.96% | 0.012 | 87.80% | 0.00% | 0.000 | 0.00% | 0.00% | 0.040 | 4.01% | 0.71% | 0.003 | 8.19% |

Inc=Incidence, SE=Standard Error. Hypertension defined as a blood pressure equal to or above 140mmHg systolic or 90mmHg diastolic.

### Predictors of hypertension cascade progression

Table 3 reports factors associated with having hypertension diagnosed (Table 3). Survey year was not associated with a change in rate of diagnosis rates. Women were 2·81 (95%CI: 1·56, 5·06) times more likely to be diagnosed than their male counterparts. Asian/Indian South Africans were 0·41 (CI: 0·21, 0·80) times more likely to be diagnosed in comparison to Africans, while White South Africans were 1·59 (95% CI: 0·47, 5·44) times more likely to be diagnosed. Increased age was associated with higher rates of diagnosis, with those aged 60-74 years being 1·24 (95% CI: 0·70, 2·19) times more likely to be diagnosed compared to those aged 35-44 years.

**Table 3.**
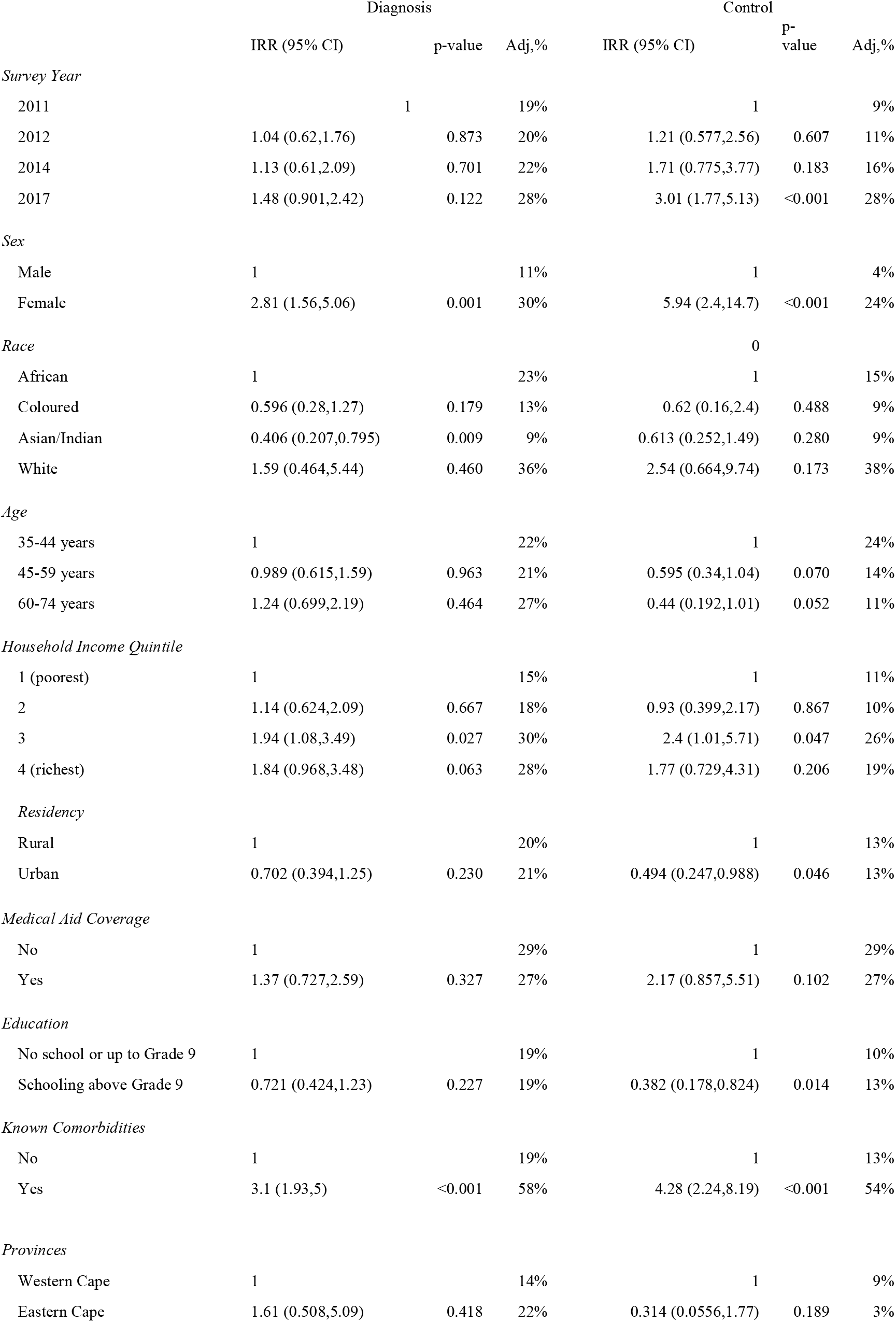
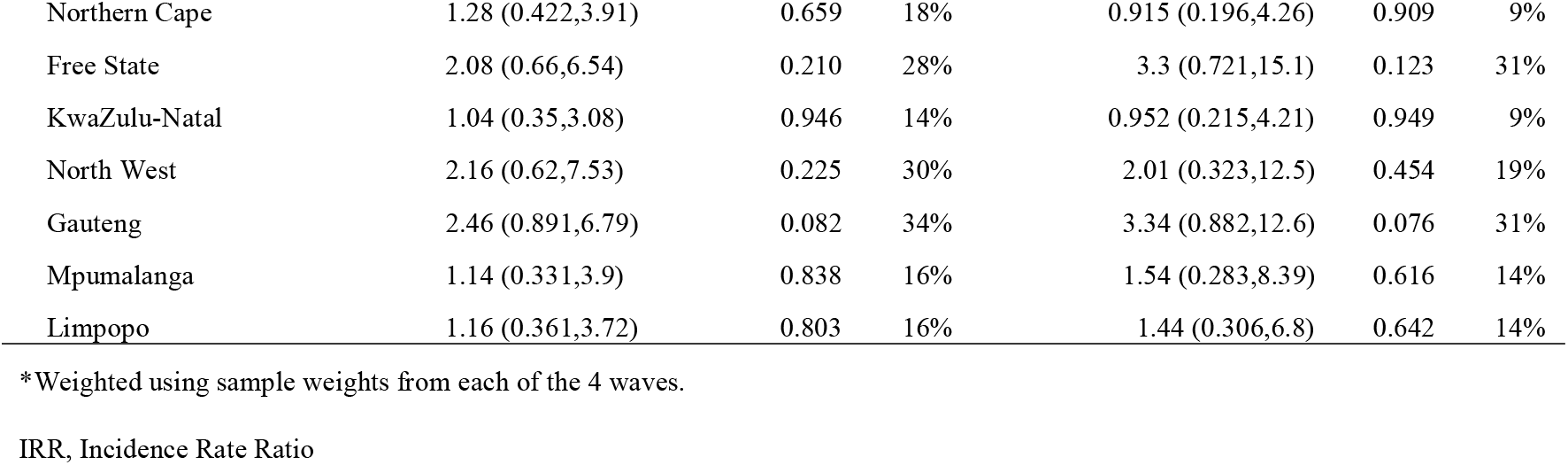
Predictors of incident hypertension control in South Africa, 2008-2017

Multiple factors were found to be important in hypertension control (Table 3). Participants who became newly hypertensive between 2014 and 2017 (wave five) had 3·01 (CI: 1·77, 5·13) times the incidence rate of being controlled compared to those who became newly hypertensive between 2008 and 2011 (wave two). Women were more likely than men to have a controlled blood pressure (IRR: 5·94, 95% CI: 2·40, 14·7) compared to men in our incident cohorts. White participants (IRR: 2·54, 95% CI: 0·66, 9·74) had higher rates of being controlled compared to African participants. Among those with incident hypertension, those aged 60-74 years had 0·44 (95% CI: 0·19, 1·01) times the incidence rate of being controlled compared to those aged 35-44 years. Finally, survey year was also found to be associated with a higher predictive probability of having a controlled blood pressure, increasing from 9·18 (95% CI: 4·91, 13·5) in survey year 2010, to 11·2 (95% CI: 5·42, 16·9) in 2012, 15·7 (95% CI: 5·68, 25·7) in 2014 and 27·6 (95% CI: 16·0, 39·3) in 2017.

## Discussion

Using the novel method of incident care cascades, we found that the proportion of people with hypertension who successfully progressed to controlled blood pressure tripled between 2010 and 2017 in South Africa. There remained a very high burden of unmet need, however, with the absolute rate of control—a measure of “met” need—reaching only 22%.

Our finding of the greatest losses in the incident hypertension care cascades coming before diagnosis is consistent with other literature from sub-Saharan Africa, which reports that the biggest unmet burden of need lies between developing disease and diagnosis.[8,26–29] Using data from the South African National Health and Nutrition Survey (SANHANES 2011-2012), Berry et al. found that 71.8% of South Africans with hypertension did not reach the diagnosis phase of the cascade, while Ware et al found that 58% of hypertensive South Africans were unaware of their diagnosis.[28,30] There are many reasons for this loss in South Africa and other low-middle income settings, such as the fact that individuals rarely seek health care for asymptomatic conditions like hypertension and that men in particular show poor health seeking behavior.[7] The SANHANES found that the mean duration since health care was last received among South Africans to be between 1·6 and 2·2 years, with 20·7% of women and 27·5% of men reporting never have sought access to public health care.[31]

Encouragingly, we found that control of hypertension in our incident cascades improved over time, from 7·1% in wave two to 22·1% in wave five. One reason might be an expanding effort in South Africa to leverage the robust HIV infrastructure to provide primary care for other conditions.[12,32] Primary care programs for HIV have been shown to improve NCD markers for quality care in multiple settings across sub-Saharan Africa, with an undetectable viral load associated with a lower systolic blood pressure and patients on ART having improved rates of progression through various phases of NCD cascades.[33,34] With the adoption of universal HIV treatment in South Africa in 2016, there have been substantial increases in HIV treatment uptake and viral load suppression, suggesting that healthcare infrastructure has been able to scale up care delivery that can also be directed towards diseases like hypertension.[35] We should note, however, that improvements in the HIV cascade do not necessarily translate to better NCD care. Muddu et al. found that in Uganda, while 90·3% of HIV patients progressed to viral suppression, only 24·3% of the same patient population had a controlled blood pressure. [26] More explicit intervention may be needed to utilize HIV infrastructure to routinely screen people for hypertension.

In addition to benefiting from HIV-related infrastructure, improvement in hypertension control may also be related to NCD-focused initiatives by the South African National Department of Health (NDoH).[11] These include the adoption of the integrated chronic disease management (ICDM) model, that included facility reorganization, outreach teams for assisted self-management, and health promotion.[32] Evaluation of the 2011 ICDM pilot through a controlled interrupted time-series analysis revealed modest gains, with only a 7·0% greater probability of having controlled blood pressure among patients in ICDM facilities, compared to routine-care facilities, possibly due to poor referrals, poor defaulter tracing, and problems with prepacking of medications, clinic appointments and patient waiting times.[12,32] The Ideal Clinic Programme, launched in 2013, aimed to address some of these concerns by establishing standards for infrastructure, adequate staff, adequate medicine and supplies, and administrative processes.[11,16] The South African government has also adopted policy level changes with the *Strategic Plan for the Prevention and Control of Non-Communicable Diseases 2013-2017*.[10] Results of this strategy included legislation that limited excessive salt and sugar intake, safe green spaces with exercise equipment to encourage physical activity and national guidelines on the management of hypertension that advocate strongly for lifestyle modification as a first line of treatment.[10,36–38] Some of these macro-level changes may have led to improved control of hypertension over time, as our analysis found. Whatever the causes, the WHO NCD Country profile for South Africa for 2017 shows that there have been substantial strides towards providing continuous access to anti-hypertensive medications at primary health care facilities.[39] Our results both confirm the value of the policy and practice revisions and underscore the large challenge that remains.

Given the current COVID-19 pandemic, it is possible that some of the progress made over recent years may be lost due to disruptions in routine care.[40] According to a WHO survey completed by 155 countries in May 2020, more than half of the countries surveyed reported disruptions in service delivery for hypertension, mainly due to staff being reassigned to support expanding health system efforts to treat COVID-19.[41] Other common reasons for disruptions were decreased availability of public transportation and the cancellation of treatments planned prior to the pandemic.[41] The long-term effects on the care cascade in South Africa remain to be seen.

We encountered several limitations in our study. Diagnosis and treatment were self-reported, and patients on medications for hypertension may be more likely to report diagnosis than those not on medications. We were not able to account for lifestyle modification in our treatment variable or assess the outcomes for people who were excluded due to missing variables. Survey effects would be of concern in a prevalent cascade, as interviewers would direct patients to care if their blood pressure were found to elevated as part of the study, thereby possibly influencing participant’ health seeking behaviour and how they progress through the cascade. Our incident cascade mitigates it by only considering newly hypertensive cases. Finally, our incident cohorts differ in hypertension incidence, signalling that our incident populations may not be entirely comparable or that our finding of improvement over time may be partly attributable to the change in composition of the sample. We aimed to mitigate this by controlling for comorbidities in the regression so that we are controlling for any differences in baseline health status in the cohort.

## Conclusion

In conclusion, the novel use of incident cohorts in this study allowed the preservation of longitudinal data for analysis, thereby allowing us to give a more accurate depiction of current system performance by negating effects of longstanding prevalent cases. We found that the proportion of people with controlled hypertension increased over time, but that there still exists a large burden of unmet need for diagnosis, treatment and control. Improvement in controlled hypertension over the study time period might be due to the South African National Department of Health moving towards a more integrated approach in the management of HIV and NCD’s through their Ideal Clinic program. Identifying key leverage points within the Ideal Clinic program may thus offer a promising avenue for further investigation to inform policy changes in similar settings.

## Data Availability

This study uses data from the first through the fifth waves of the National Income Dynamics Study (NIDS). NIDS is a nationally representative panel survey that was implemented by the South African Labour and Development Research Unit at the University of Cape Town. The data used in the present study are publicly available and can be accessed at the following link: http://www.nids.uct.ac.za/ Data for all 5 waves of the NIDS were accessed using the DataFirst portal following registration. Ethics approval for NIDS data collection for Waves 1 to 5 were granted by University of Cape Town's Commerce Faculty Ethics in Research Committee. For Wave 5 additional ethics approval was granted by University of Cape Town's Faculty of Health Sciences Human Research Committee.

---

**Supplementary Figure 1:**
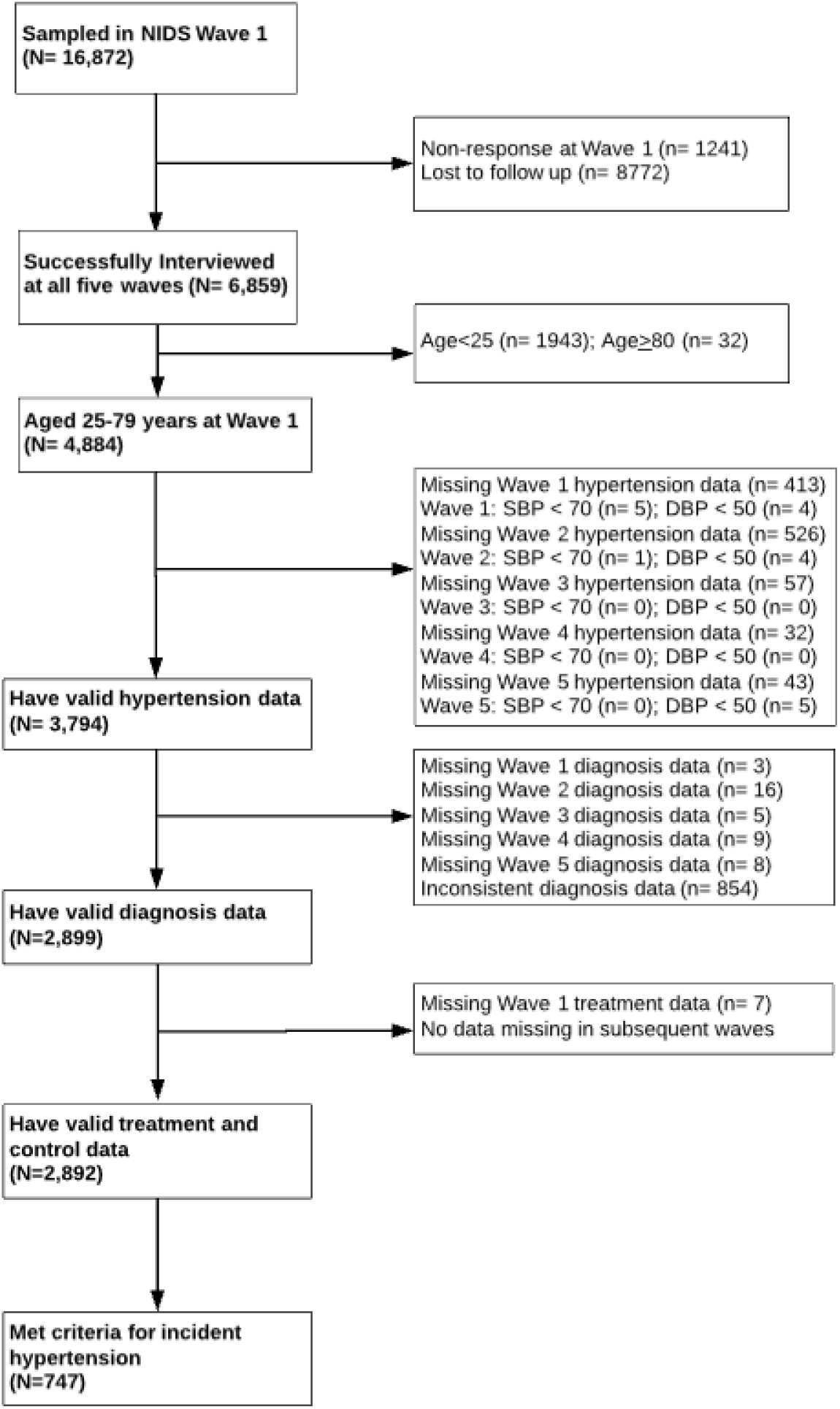
Flow diagram of exclusion criteria

**Supplementary Figure 2.**
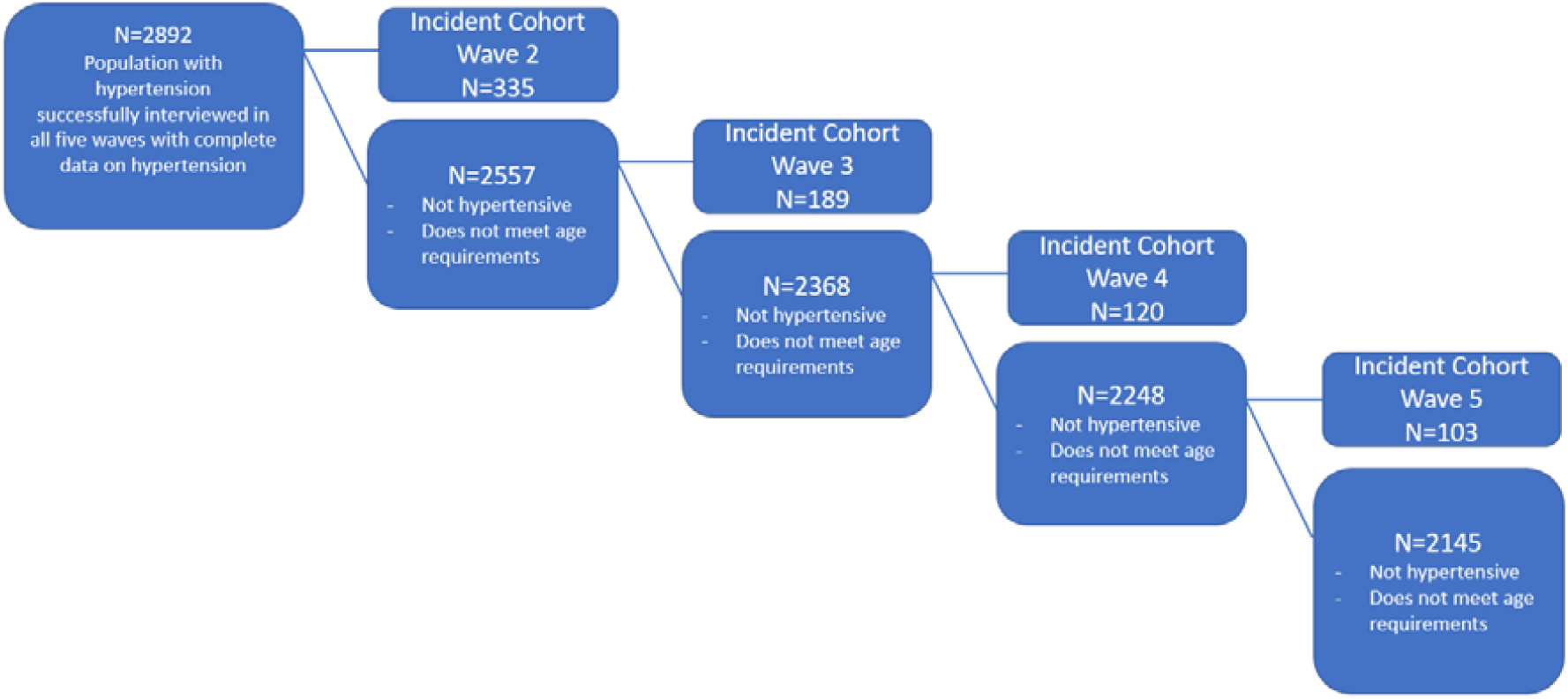
Diagram illustrating how incident cohorts were created

